# Early estimates of COVID-19 infections in small, medium and large population clusters

**DOI:** 10.1101/2020.04.07.20053421

**Authors:** AS Siraj, A Worku, K Berhane, Y Berhane, DS Siraj

## Abstract

Since its emergence in December 2019, COVID-19 has rapidly developed into a pandemic with many countries declaring emergency conditions to contain its spread. The impact of the disease, while it has been relatively low in the Sub Saharan Africa (SSA) so far, is feared to be potentially devastating given the less developed and fragmented health care system in the continent. In addition, most emergency measures practiced may not be effective due to their limited affordability as well as the communal way people in SSA live in relative isolation in clusters of large as well as smaller population centers. To address the acute need for estimates of the potential impacts of the disease once it sweeps through the region, we developed a process-based model with key parameters obtained from recent studies, taking local context into consideration. We further used the model to estimate the number of infections within a year of sustained local transmissions under a total of 216 scenarios that cover different sizes of population, urban status, effectiveness and coverage of social distancing, contact tracing and usage of cloth facemask. We showed that when implemented early, 50% coverage of contact tracing and facemask, with 33% effective social distancing policies can “flattens the curve” of local epidemics and even bending it enough to result in fewer cumulative infections, bringing the pandemic to a manageable level for all population sizes we assessed. In SSA countries with limited healthcare workforce, hospital resources and ICU care, a robust system of social distancing, contact tracing and facemask use could yield in outcomes that prevent several millions of infections and thousands of deaths across the continent.

**Funding:** No funding source.

## Introduction

Since its emergence in December of 2019, SARS-CoV-2 virus has continued to spread in many regions with 3.2million confirmed cases and more than 228 thousand reported deaths worldwide in more than 210 countries and territories as of April 30, 2020.[1] The rapid progression of the number of infections and deaths due to SARS-CoV-2 has taken many by surprise, with many of its characteristics related to its transmissibility under continuous update.[2] While the disease is still actively spreading in many regions of the world, researchers are working to quantify transmission parameters [2–7], and make estimates of infections and resulting deaths under different scenarios [8–11]. However, these studies are either too specific to certain geographies [10,12] or are too general to handle realistic scenarios [8,11] in the context of Sub Saharan Africa where majority of the population live in rural and semi urban areas with weak physical interconnections, and urban areas with limited access to basic water and sanitation facilities and space to self-isolate. To overcome this problem and make estimates of infections once sustained local transmission occurs in clusters of population we chose three urban scenarios with 3 million, 1 million and 100 thousand population sizes and one rural setting with population of 200 thousand. we developed a process-based model structured into Susceptible-Exposed-Infectious-Removed (SEIR) compartments, with most parameters obtained from recent studies, and based on 216 scenarios of initial number of cases, population sizes, effectiveness of social distancing, coverage of contact tracing and facemask, more suited to realities in resource poor, less interconnected regions in SSA.

## Methods

We developed a process-based model with four human compartments: Susceptible (S), Exposed (E), Infectious (I), and Removed (R) [13,14]. Because estimates of the basic reproduction number *R_0_* and length of the infectious period cannot be viewed in isolation, we used estimates in their proper context and paired *R_0_* and length of infectious period estimates from similar sources [7,12]. In our model setup, contact tracing is assumed to affect those that are in the infectious state resulting in their transition to the removed state *R*. Social distancing reduces the contact rate between individuals. At the same time, use of facemask reduces the probability of infection given contact. Based on effectiveness of medical facemask estimated at 41% [15], we assumed the use of population level facemasks made of cloth will have 25% effectiveness.

Initially, we tested the model for the four locations and one additional small rural location with population of 100k. However, we dropped the small rural settings as most results were similar to those under the larger rural settings we have included. All model simulations were performed using the Partially Observed Markov Process (Pomp) package [16], available in R, with all scenarios run based on parameters either fixed or Monte Carlo sampled from their corresponding distributions. We run all scenarios for 100 realizations to account for stochasticity in the model process and to quantify uncertainties in the parameters. Results are reported as median effect estimates of new infections and 95% credible intervals (CI) on a daily scale.

### Role of the funding source

There was no funder for the study. The corresponding author had full access to all the data in the study and had final responsibility for the decision to submit for publication.

## Results

### Counterfactual scenario

Our results show that with no interventions of social distancing, contact tracing and facemask usage, transmission would result in 94% and 80% of the population infected over a year’s period in urban and rural settings, respectively. In addition, nearly all infections in the urban settings and more than two-third of the infection in the rural settings would happen in the initial 90 days after the trigger point of community level transmission (Figure 1, left panel).

**Figure 1:**
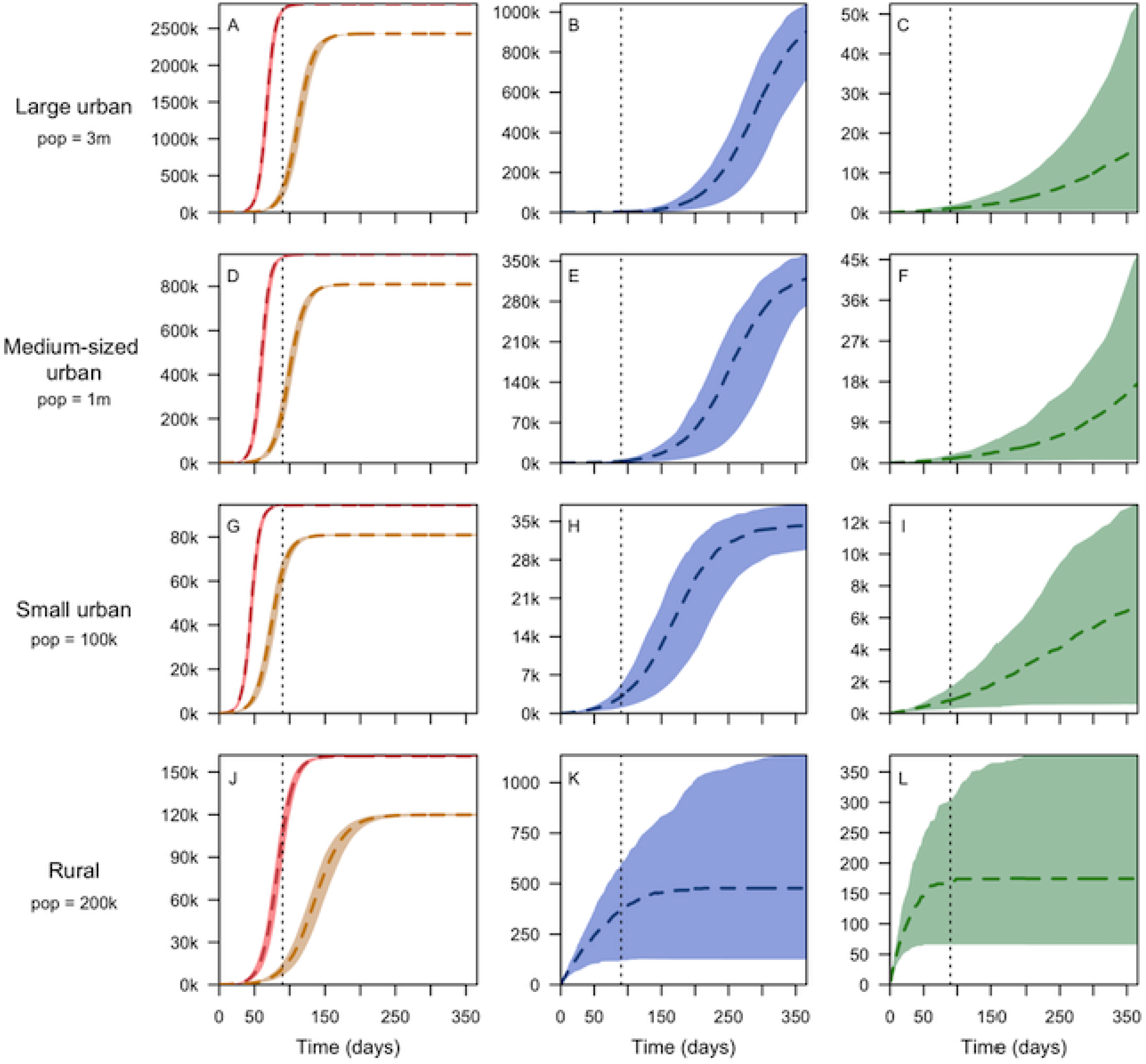
Cumulative number of infections in one year after the trigger point (50 initial infections) in large urban (A-C), medium sized urban (D-F), small urban (G-I), and rural (J-L)settings from top to bottom respectively, assuming no social distancing (left panel – red), with social distancing (33% in urban and 25% in rural) only (left panel-brown), with social distancing and 50% contact tracing (middle panel); and with 50% coverage of both contact tracing facemask use (right panel). The short-term projections (within the first 90 days) are shown by the dotted vertical lines. Curves (broken lines) show the median values and corresponding shaded regions show the 95% credible intervals.

### Effects of social distancing

In the absence of other interventions, the effects of 33% and 25% social distancing in urban and rural setting were minimal in the long-term, with only 14% (95% CI 13.7%-15%) and 26% (95% CI 24.1%-27.3%) reduction in the number of infections over one-year period respectively. In general, these reductions are small since the base projection (non-intervention) concerns large proportion of the population. In the short-term, however, these relative reductions increase to 88.4% (95% CI 83%-93%), 72% (95% CI 64%-80%), 30% (95% CI 25%-38%), and 89% (95% CI 81%-94%) in large urban, medium-sized urban, small urban, and rural settings respectively (Figure 1, left panel).

### Added effect of social distancing and contact tracing

Based on our model projections, the effects of contact tracing are significantly large for all geographic settings when coupled with social distancing with effectiveness of 33% and 25% in urban and rural settings, respectively. Our results show additional decrease in the number of infections of 54% (95% CI 49%-62%) 52% (95% CI 47%-57%), 49% (95% CI 45%-54%), and 74% (95% CI 73%-75%) in large urban, medium-sized urban, small urban, and rural settings respectively (Figure 1, 2^nd^ column; Figure 2C). This dramatic decrease amounts to 60% (95%CI 53-73%) and 99% (99-100%) reduction in infections in urban and rural settings respectively from those considering 33% and 25% social distancing only. The results in the rural setting indicate social distancing and contact tracing can lower the epidemic to manageable levels (Figure 3). The added effect of contact tracing is not only to lower the number of infections but also to delay the peak transmission period towards the end of the year, effectively flattening the curve (Figure 2C).

**Figure 2:**
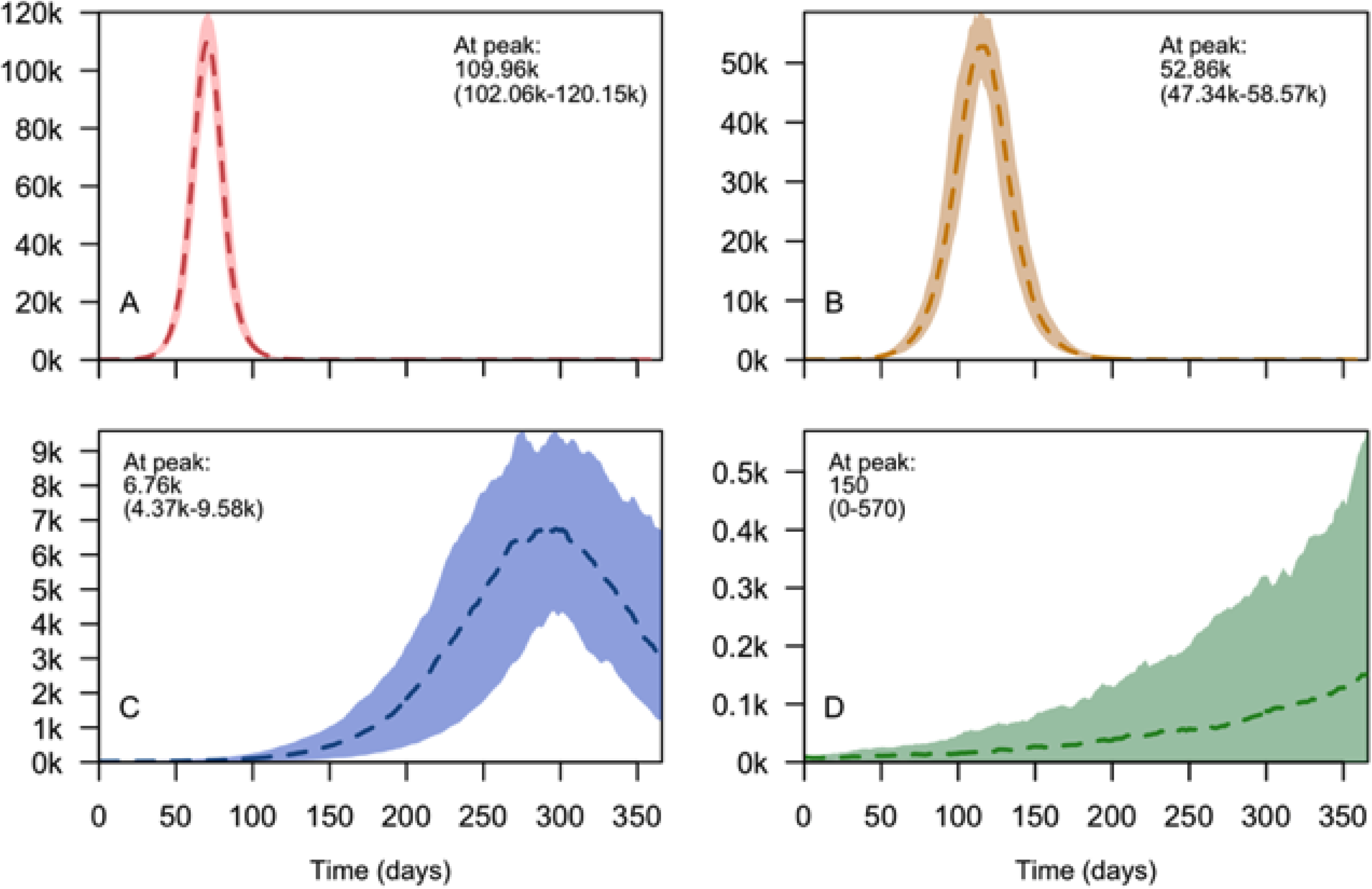
Daily number of infections in a large city of 3 million population in with different levels of interventions. Figures show infections with no interventions assumed (A), 33% social distancing assumed (B), 33% social distancing and 50% contact tracing assumed (C), and 33% social distancing, 50% contact tracing and 50% facemask use assumed (D). Curves (broken line) show the median value and corresponding shaded regions show the 95% credible interval.

**Figure 3:**
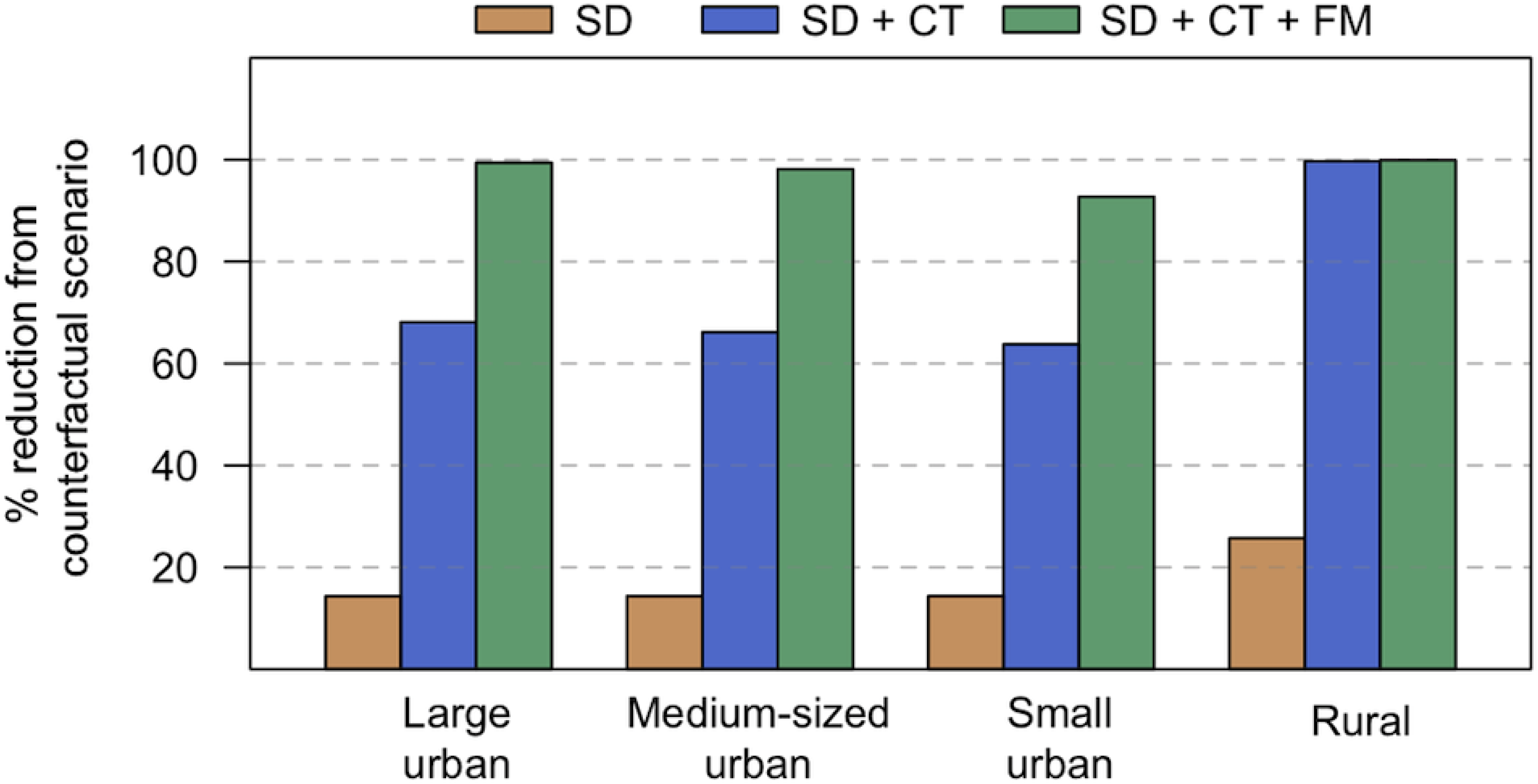
Median percentage reduction of infections from the counter-factual scenario as a result of the three interventions serially stacked up on each other. The interventions included are social distancing (33% in urban and 25% in rural) only (brown), social distancing and 50% contact tracing (blue); social distancing and 50% coverage of both contact tracing and facemask use (green). The small and medium-sized urban settings have relatively smaller overall infections averted due to the effect of our frequency dependent dynamics leading to proportionally larger population getting infection in areas with smaller population.

### Added effect of social distancing, contact tracing and facemask usage

When all three non-pharmaceutical interventions are applied, with 33% and 25% social distancing in urban and rural settings, 50% contact tracing, and 50% facemask use, our results show a large number of infections could be averted especially in urban settings. Accordingly, these interventions would bring the number of infections down by additional 31% (95% CI 22%-37%), 32% (95% CI 24%-38%), 29% (95% CI 18%-40%), and 0% (95% CI 0%-1%) in large urban, medium-sized urban, small urban, and rural settings respectively (Figure 1, 3^rd^ column, Figure 2D).

The added effect of facemask is minimal in the rural setting because all infections would have been averted by social distancing and contact tracing only (Figure 3). The results also show the added benefit of facemask in delaying the peak transmission period further in time (Figure 2D). The overall effect of all three interventions is relatively low in small urban settings due to the effect of frequency – dependent disease dynamics (Figure 3). In such dynamics, equal number of initial cases in different sized population will lead to similar number of infections for some time, thus will have large proportion of infected population in the small-sized communities.

### Effects of early intervention

Rolling out early action with the implementation of social distancing, contact tracing and facemask could help avoid rapid growth of the epidemic and mitigate problems related to capacity of healthcare delivery. Compared to a scenario that triggers action after an initial 100 cases, our results show early action triggered by 50 initial cases lowered the daily numbers of infections approximately by half over the initial 90 days (Figure 4B-D). This indicates early action (when there were 50 infection in the community) rather than late action (when there were 100 infections) would bring the daily number of cases by half throughout the initial epidemic growth, allowing health facilities to respond better to patients with mild and sever manifestations of SARS-Cov-2 infections.

**Figure 4:**
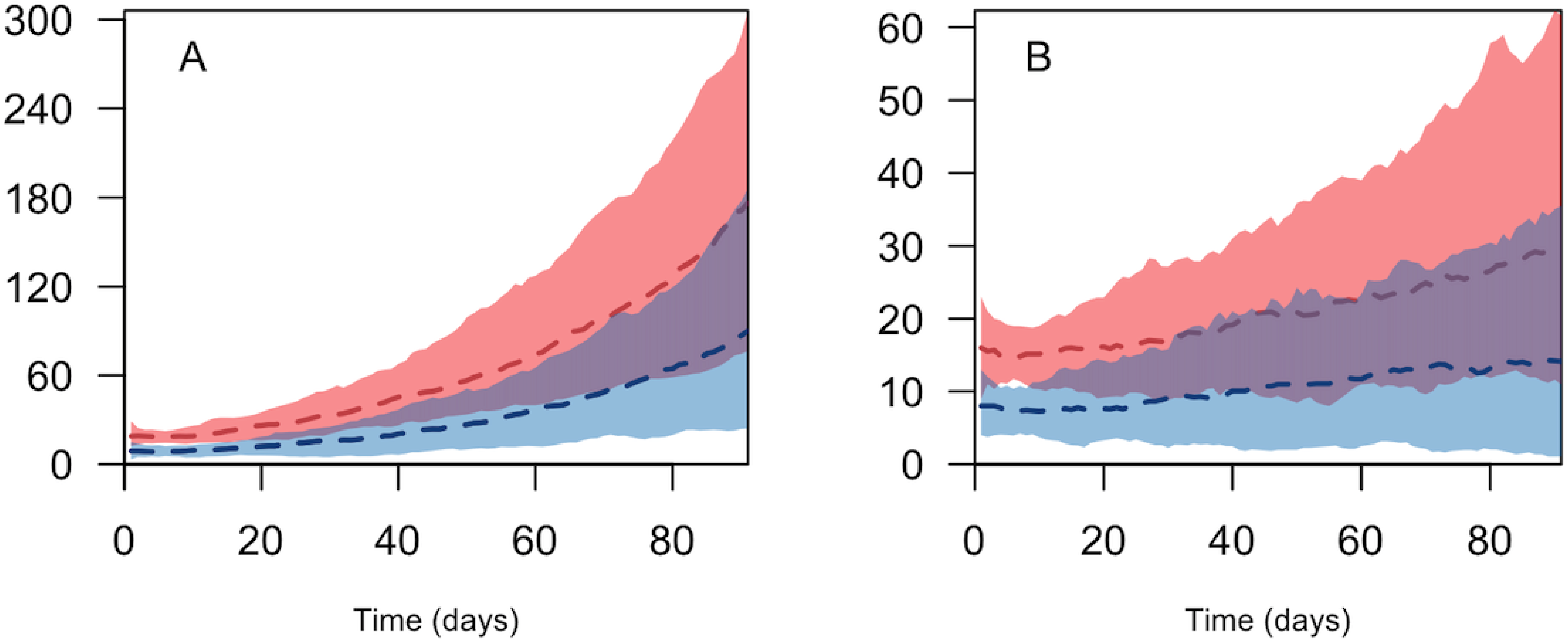
Daily number of infections in a large city of 3 million population in the initial 90 days and initial infections of 100 (red) and 50 (blue), with 33% social distancing and 50% contact tracing assumed (A), and 33% social distancing, 50% contact tracing and 50% facemask use assumed (B). Curves (broken line) show the median value and corresponding shaded regions show the 95% credible interval. Earlier action (beginning when 50 infections occurred) would lead to reduction of infections by half in both scenarios early in the epidemic.

### Effects of facemask and social distancing measures

Easing of social distance measures could be a difficult policy decision for many SSA countries affected by this pandemic. To quantify the effect of relaxing social distance measures, we started with a scenario where the initial social distancing effectiveness of 33% in urban 25% in rural areas are maintained in addition to having 50% contact tracing coverage but excluding facemask (Figure 5A and 5D). We then incrementally lowered the effectiveness of social distancing, while increasing facemask coverage to maintain the same level of infections over one-year period. Our results show a non-linear association between easing social-distancing and facemask coverage both in urban (Figure 5C) and rural settings (Figure 5F). Our results further showed that, for the scenario considered, decreasing social distancing from 33% to 25% in urban settings and from 25% to 15% in rural settings would require 43% and 47% coverage of facemask to make up for the increase in the number of infections as a result of decrease in social distancing respectively (Figures 5C and 5F. These figures are less than the 50% facemask coverage show in Figure 5B and 5E for urban and rural settings.

**Figure 5:**
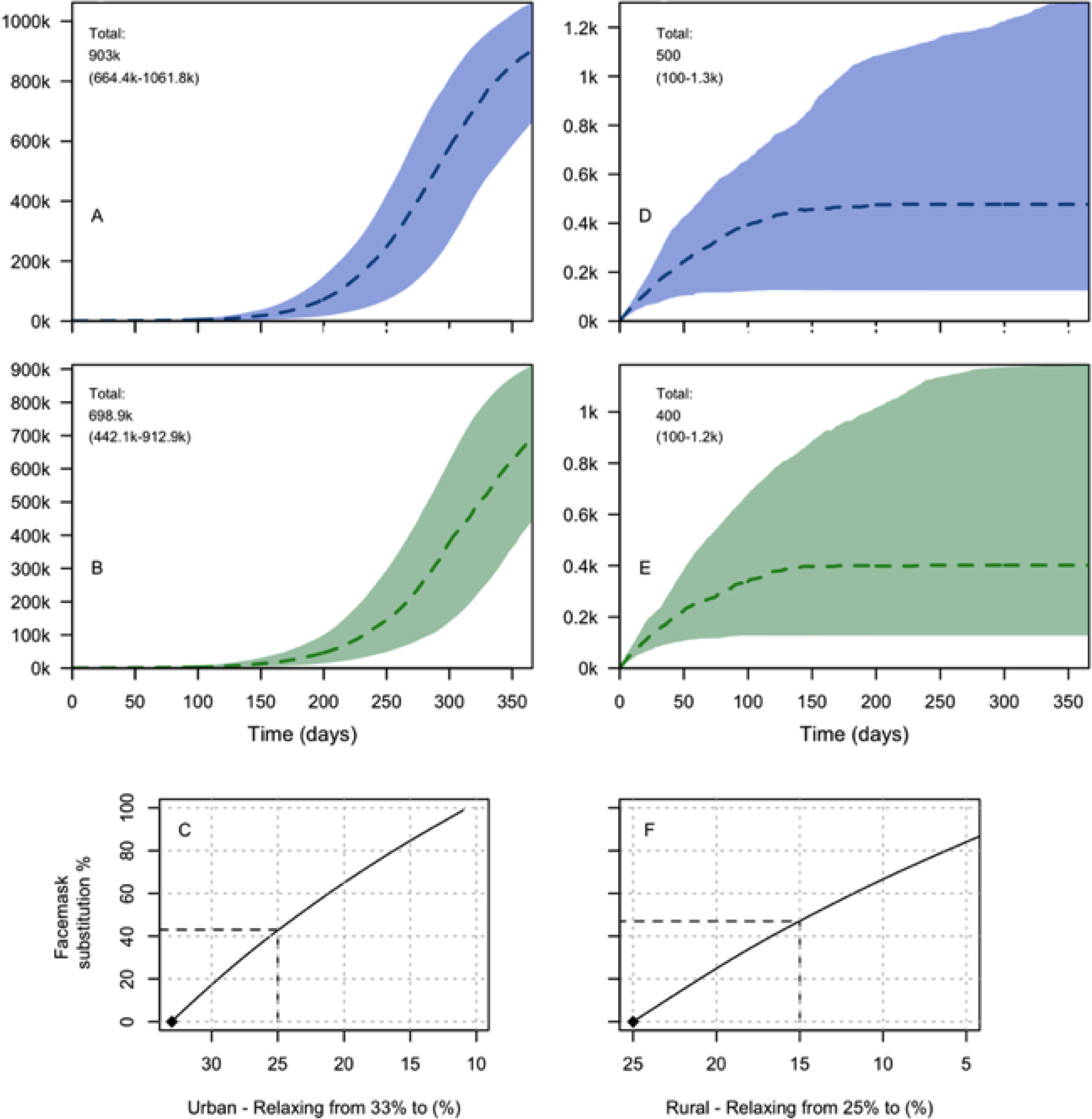
Cumulative number of infections in a large city of 3 million population with 33% social distancing and 50% contact tracing assumed (A), and after relaxing social distancing to 25% social distancing, maintaining 50% contact tracing, and added 50% facemask use (B). Cumulative number of infections in a large rural district of 200k population with 25% social distancing and 50% contact tracing assumed (D), and after relaxing social distancing to 15% social distancing, maintaining 50% contact tracing, and added 50% facemask use (E). Figures C and F show the coverage of facemask introduced to substitute the effect of relaxing social distancing in the respective urban and rural settings. Broken lines in C and F show facemask coverage that would have offset the effect of relaxing social distancing from those shown in A and D respectively.

## Discussion

We developed a stochastic process-based model with parameters from recent published works, to estimate infections within the first 90 days and one year of initial cases for scenarios more realistic to many African settings in terms of population size and capacity to carry out interventions such as lockdown and closure of ports of entry. Our estimates show effective contact coupled with social distancing and cloth facemask use could bring down the number of infections and associated deaths to manageable levels when implemented early and in combination. These results confirm recent evidences of the roles social distancing [17], contact tracing [8,11], and facemask use [18] play as reliable non-pharmaceutical intervention against SARS-Cov-2 at the initial stage of a local epidemic.

Our models have assumed that transmission of COVID-19 happens at a community level where reported cases may not have any obvious imported sources. By the time such a case is reported and diagnosed, due to delays in reporting and testing, we assumed that there could be at least 50 infections going around unobserved. This is because even with effective contact tracing and isolation in place, due to possible delays in locating suspects, secondary and tertiary infections may have occurred without being traced and quarantined. We tested our model by considering lower number of initial infections (1< I < 10), which resulted in lower probabilities of an epidemic growth due to the effect of contact tracing and stochasticity in our model leading to zero infections after sometime (not shown).

With the virus rapidly spreading around the world, Sub-Saharan Africa is anticipating a start of epidemic levels in disease and death rates as seen in other regions, with only 38,314 confirmed cases and 1,595 deaths reported in 42 countries as of April 30, 2020 [1]. Though each African nation’s situation may be unique, patterns of population settlement and physical connectedness, as well as living conditions in most countries can be generalized into one of isolated rural, semirural or large urban settlements, the latter with a possibility of having slum area. We contend our classifications of population centers into three urban locations with population of 100 thousand, one million and three million and two rural districts with population of 100 thousand and 200 thousand represents the reality in most countries in SSA. To decide which scenario best representatives their situation, each country should consider their status of community transmission of SARS-CoV-2, strength of social distancing measures taken, risk of imported new cases, number of confirmed cases, the strength of the field epidemiology work to trace contacts and isolate, presence of isolation facilities, coverage of facemask usage[15], and population density and sizes. In population centers where the potential for becoming new epicenter of transmission is high, these measures should be accompanied by travel restrictions between clusters to prevent importation of cases.

The effect of increased contract tracing and isolation is not only lowering the total attack rate and mortality, but also delaying the time to the peak transmission slowing build-up of cases, thus allowing hospitals and healthcare systems mobilize their workforce and other resources to effectively deal with this devastating emergency. This so-called “flattening the curve” scenario [13] will allow critical resources, including health care workers, hospital beds, and medical equipment and supplies, to be utilized efficiently leading to more lives saved.

Sub Saharan Africa’s large young population is expected to have relatively lower mortality rates given that the mortality rates so far observed was highest among older population [11]. In addition, the majority of the population in SSA live in rural areas where the overall population density is low making natural physical distancing easier, thus mitigating the impact of the SARS-CoV-2 pandemic [7,13]. Accordingly, with the age specific fatality rates from China [19] based on age pyramid obtained from Ethiopia [20], our results for the scenario of 33% social distancing, 50% contact tracing and 50% facemask use would have averted 12.1k (95% CI 11.9k-12.2k) deaths in the large urban setting.

On the other hand, weak health systems, poor access to sanitation, and overcrowded cities and slums that make social distancing a challenge can negatively impact the spread of the virus. A widespread use of cloth facemask/ cover in urban areas, where people may find it hard to keep distances, thus can alleviate the challenge of social distancing needed to decrease transmissibility of SARS-CoV-2 [21] especially in urban centers where people may find it hard to keep distances. We have modeled our estimate based on three basic reproduction number *R_0_* values, three cluster sizes and three initial cases. SSA countries and regions are advised to take their situation into account [22] when they use estimates from these scenarios.

The large differences in the number of infected individuals between scenarios initialized with 50 cases and 100 cases (Fig 4) demonstrate the effect of early action preventing large transmissions and lowering the size of the epidemic. This suggests a need for countries to quickly undertake extensive training of the public health workforce to identify and isolate contacts of those who are diagnosed with SARS-CoV-2. Currently, Africa has the lowest testing rate of any part of the world [23]. Since testing is a necessary precondition for contact tracing and isolation, countries should scale up their testing, contact tracing and isolation capacities within the short window of opportunity that remains to minimize the local impact of the pandemic.

Our model is not without limitations. We have not considered symptomatic and asymptomatic infections separately, which may have overestimated the force of infection due to the lower shedding probability among non-symptomatic COVID-19 cases [12]. We also accounted for contract tracing by considering a fraction of the incubation period, unlike others who have used more realistic distribution-based time to isolation [8]. We have assumed that contact tracing will be completely effective with no onward transmission from those individuals suspected of the disease, which may not be realistic given the less optimal process in identifying suspects, and putting them in safe isolated location in many African settings. The estimates that we provided for mortality should also be taken only as rough initial attempts, as not much is known about the case fatality rate in SSA and also potentially higher levels of mortality due to lower levels of immunity in the populations (even if young) and impacts of underlying health conditions. Finally, the fact that our model does not have age structure has limited our capacity to make more realistic estimates of age specific mortality.

## Data Availability

This paper does not used data apart from model parameters.

## Acknowledgments

We would like to thank Abigail Maps for her help in editorial works to make the manuscript more readable. We also thank Dr. Demissie Alemayehu and Dr. Maru Aregawi for their comments and suggestions in the early stage of the manuscript.

## Notes

### Competing Interest Statement

The authors have declared no competing interest.

### Funding Statement

No external funding was received for this study.

